# Association between fractional exhaled nitric oxide (FeNO) and peripheral eosinophils in patients with asthma: a retrospective cohort study

**DOI:** 10.1101/2025.02.15.25322353

**Authors:** Yahya Z. Habis, Abdullah Sakkat, Shahad H. Alaidaroos, Reema D. AlGhamdi, Nada H. Almuwalad, Joud M. Aljuhani, Atheer A. Alharbi

## Abstract

**Background:** Fractional exhaled nitric oxide (FeNO) and peripheral eosinophils are important biomarkers of Type 2 inflammation in asthma. FeNO reflects airway inflammation, while eosinophil counts indicate systemic eosinophilic activity. Their combined utility in asthma management warrants further investigation.

**Objective:** This study aimed to assess and compare FeNO levels and peripheral eosinophils in asthmatic patients, examining their relationship with disease characteristics and bronchodilator responsiveness (BDR).

**Methods:** A retrospective analysis of 101 adult asthmatic patients at King Abdulaziz University, Jeddah, was conducted. Patient data on FeNO, eosinophil counts, spirometry, and comorbidities were collected and analyzed using correlation and receiver operating characteristic (ROC) curve analysis.

**Results:** The mean FeNO level was 68.9±61.3 ppb, and the mean eosinophil count was 310.5±344.2 cells/μL. FeNO levels were significantly higher in patients with eosinophil counts ≥300 cells/μL (p=0.012) and correlated positively with BDR (r=0.38, p=0.001) but not with eosinophil counts (r=0.16, p=0.11) or asthma control test (ACT) scores. Eosinophil counts were significantly associated with moderate airway obstruction (p=0.041). FeNO showed a borderline ability to differentiate allergic rhinitis (p=0.059) but limited performance in identifying other comorbidities, such as chronic rhinosinusitis (p=0.17), GERD (p=0.87), and eczema/atopic dermatitis (p=0.11 and p=0.24, respectively).

**Conclusion:** FeNO and peripheral eosinophil counts are valuable complementary biomarkers for assessing type 2 inflammation in asthma and guiding treatment. However, their variability across clinical contexts underscores the need for integration with other assessments. These findings support a multidimensional approach to asthma management and call for further research to refine their clinical applications.

## Introduction

Asthma is the commonest chronic lung disease with airway inflammation presenting a central component, but clinical symptoms and standard spirometry inadequately reflect the underlying inflammatory processes [1]. Airway remodeling in severe asthma is considered irreversible; yet its components as a cause of clinical symptoms and/or lung function changes stay unidentified [2]. Asthma control is assessed according to the manifestations, lung function as well as the type and extent of airway inflammation [3]. Achieving remission in asthma is a key treatment goal; however, there is still no consensus on its definition despite extensive multidisciplinary efforts [4]. The appearance of eosinophils in airways presents a dynamic integration of different cell types within lung tissue and does not reflect a passive effect of inflammation. The cellular crosstalk leads to continuation of eosinophilic inflammation, exacerbation and long-term damage to the airway structure and function [5].

There is a considerable increase in the occurrence of bronchial asthma in the Kingdom of Saudi Arabia [6]. The Global Burden of Disease study also revealed that the Middle East and Africa (MEA) has the highest incidence of asthma in young adults, following South Asia and North America in 2019; the largest changes in incidence rates were observed in countries such as Saudi Arabia, Oman, and Sudan [7].

Type 2 inflammation is a prominent feature of asthma with elevated eosinophil count and several mediators forming an attractive therapeutic target [8]. Sputum inflammatory cell analysis is the gold standard in discriminating eosinophilic inflammation in asthma yet with restrictions in its clinical applicability and may be influenced by factors such as steroid intake, comorbidities and environmental exposures or habits [9]. It is thus not considered in routine practice due to its heavy time- and staff demands [10]. In asthma, exhaled nitric oxide (NO), produced in response to inflammatory cytokines and mediators in the airways, is easily identified thus, serving as a promising marker for disease monitoring [11]. The scientific community is faced with limits in the use of asthma biomarkers. Accordingly, there is growing interest in peripheral eosinophilia and fractional exhaled nitric oxide (FeNO) to better reflect the condition [9].

The FeNO is a reliable, readily available and feasible test recommended in asthmatic adults and children with a key role in management [11]. It originates primarily in the bronchial epithelium and is produced in large quantities by the enzyme inducible nitric oxide synthase (iNOS) [12]. In asthma, FeNO is increased in eosinophilic-mediated inflammatory pathways and reflects type 2 inflammation. It is considered a key biomarker in the diagnostic process and in predicting encouraging response to inhaled steroids and biologic treatments [11]. On going research is being conducted to explore the clinical applications of FeNO measurements in other respiratory diseases, such as COPD and interstitial lung disease [13].

The aim of the present work was to assess and compare the FeNO level and peripheral eosinophils in patients with asthma. Considering the relation to the disease characteristics and bronchodilator responsiveness (BDR).

## Patients and methods

This retrospective study included 101 patients with confirmed asthma presenting to the pulmonology unit, Internal medicine Department, King Abdulaziz University, Jeddah, Saudi Arabia. Data collection took place between December 2024 and January 2025, reviewing medical records from January 2023 to July 2024. Patients were identified using the ICD codes and manual chart review. The study was approved by the ethical committee of King Abdulaziz University (HA-02-J-008) and was in accordance with the principles of the Declaration of Helsinki. Informed consents are provided by all patients.

Patients’ charts were retrospectively reviewed and data regarding the age, gender, body mass index (BMI), smoking status, presence of allergic rhinitis, chronic rhinosinusitis with nasal polyps, gastroesophageal reflux disease (GERD), and eczema/atopic dermatitis were collected using standardized data collection sheet according to the following inclusion and exclusion criteria. *Inclusion criteria*: Adult patients (>18 years) who have been diagnosed with asthma and did FeNO test. *Exclusion criteria*: Patients < 18 years, non-asthmatic, did not do FeNO test or without an available complete blood count (CBC) with differential.

Clinical evaluation was performed to all patients and data on the presence of type 2 inflammation diseases were recorded. Asthma Control Test (ACT) [14] as well as bronchodilator responsiveness (BDR) were assessed. Spirometry was measured including forced expiratory volume (FEV_1_) pre and post bronchodilator (BD). Airway obstruction post-BD was graded according to the American Thoracic Society and European Respiratory Society (ATS/ERS) guidelines as mild (FEV1% ≥70%), moderate (FEV1% 60-69%), moderately severe (FEV1% 50-59%), severe (FEV1% 35-49%) and very severe (FEV1% <35%) [15].

Peripheral eosinophil counts and percentages were reported. Counts at a cut-off ≥150 cells/μL were diagnostic of type 2 inflammation as per global initiative of asthma (GINA) and ≥300 cells/μL were highly significant while, counts <150 cells/μL was considered low (non-type 2 inflammation) [16].

The FeNO was assessed and graded (High >50, Intermediate 25-50, Low < 25) according to the ATS/ERS criteria [17].

### Statistical analysis

Statistical Package for Social Science (SPSS) program version 25 was used for analysis of data. Data was presented as number, and frequency (%) or mean±SD, range. Mann-Whitney test was used for analysis of two quantitative non-normally distributed data. Spearman’s correlation test was used. Receiver operating characteristic (ROC) and area under the curve (AUC) was plotted to determine the predictive potential of FeNO to detect type 2 inflammation disease. Results were adapted for missing variables. Significance was set at p <0.05.

## Results

The study included 101 asthmatic patients with a mean age of 46.6±14.6 years and were 70 females and 31 males (F:M 2.3:1). Characteristics of the patients are presented in table 1. 19 patients were of normal BMI, 6 were underweight, 29 were overweight and 47 were obese.

**Table 1.** Characteristics of asthmatic patients.

| Parameter | Asthmatic cases (n=101) |
| --- | --- |
| Age (years) mean±SD | 46.6±14.6 |
| Female sex – no. (%) | 70 (69) |
| BMI mean±SD | 29.2±6 |
| Smoking n (%) | 25 (24.8) |
| *Co-morbidities* n (%) |  |
| Allergic rhinitis | 58 (57.4) |
| Chronic rhinosinusitis with NP | 13 (12.9) |
| GERD | 43 (42.6) |
| Eczema/Atopic dermatitis | 20 (19.8) |
| ACT – controlled n (%) | 38 (37.6) |
| Peripheral eosinophils (cells/μL) mean±SD | 310.5±344.2 |
| <150 n (%) | 37 (36.6) |
| ≥150 n (%) | 34 (33.7) |
| ≥300 n (%) | 30 (29.7) |
| Eosinophils (%) mean±SD | 4.16±4.1 |
| *Spirometry* |  |
| FEV1 % post BD (n=68) mean±SD | 80.1±18 |
| Mild (FEV1% ≥70%) n (%) | 51 (50.5) |
| Moderate (FEV1% 60-69%) n (%) | 11 (10.9) |
| Moderately severe (FEV1% 50-59%) n (%) | 3 (3) |
| Severe (FEV1% 35-49%) n (%) | 2 (2) |
| Very severe (FEV1% <35%) n (%) | 1 (1) |
| BDR (%) mean±SD | 3.8±4.4 |
| FeNO Grade *low* n (%) | 18 (17.8) |
| *Intermediate* n (%) | 37 (36.6) |
| *high* n (%) | 46 (45.5) |
| FeNO (ppb) mean±SD | 68.9±61.3 |

BMI: body mass index, NP: nasal polyposis, ACT: asthma control test, ICS: inhaled corticosteroids, LABA: long-acting $\beta$ 2 agonist, LAMA: long-acting muscarinic antagonist, FEV1: forced expiratory volume, FVC: forced vital capacity, BDR: bronchodilator responsiveness, FeNO: fractional exhaled nitric oxide.

There was no significant difference in the mean FeNO according to gender (males 70.7±70.2 vs females 68.1±57.5 ppb, p=0.86). FeNO tended to be higher in those currently or ex-smoking (n=3, 105.3±95.9 and n=11, 93±74.9 ppb) compared to non-smokers (n=87, 64.6±58 ppb) (p=0.85) and was similar according to the BMI classes (p=0.39). The mean FeNO level tended to be higher in patients with allergic rhinitis (76.7±66.8 *vs* 58.4±52 ppb, p=0.13), chronic rhinosinusitis with nasal polyposis (83.9±61.3 *vs* 66.7±61.3 ppb, p=0.36), GERD (68.9±62 *vs* 68.9±61.4 ppb, p=0.99) and eczema/ atopic dermatitis (74.7±56.3 *vs* 67.4±62.7, p=0.62).

Post-BD FEV1 results were available for 68 patients. The mean FeNO according to the grade of airway obstruction assessed by FEV1 post-BD (mild n=51, 65.9±66.3 ppb, moderate n=11, 81.6±78.5 ppb, moderately-severe n=3, 94±101.7 ppb, severe n=2, 47.5±3.5 ppb and very severe in 1 case, 17 ppb; p=0.81). The mean FeNO tended to be higher in those with controlled ACT score (n=38, 77.8±74.7 ppb) compared to those partially (51.5±28.4 ppb) and uncontrolled (68.9±45.02 ppb) (p=0.47).

The FeNO was significantly higher in those with eosinophilic count ≥300 cells/μL (n=30, 96.2±82.1 ppb) compared to those with levels <150 (n=37, 59.6±35.3 ppb) or 150-299 (n=34, 54.9±55.9 ppb) (p=0.012) (Figure 1) and tended to correlate with eosinophil absolute count (r=0.16, p=0.11). The FeNO level did not significantly correlate with the BMI (−0.04, p=0.7) but significantly correlated with BDR (r=0.38, p=0.001) (Figure 2).

**Figure 1:**
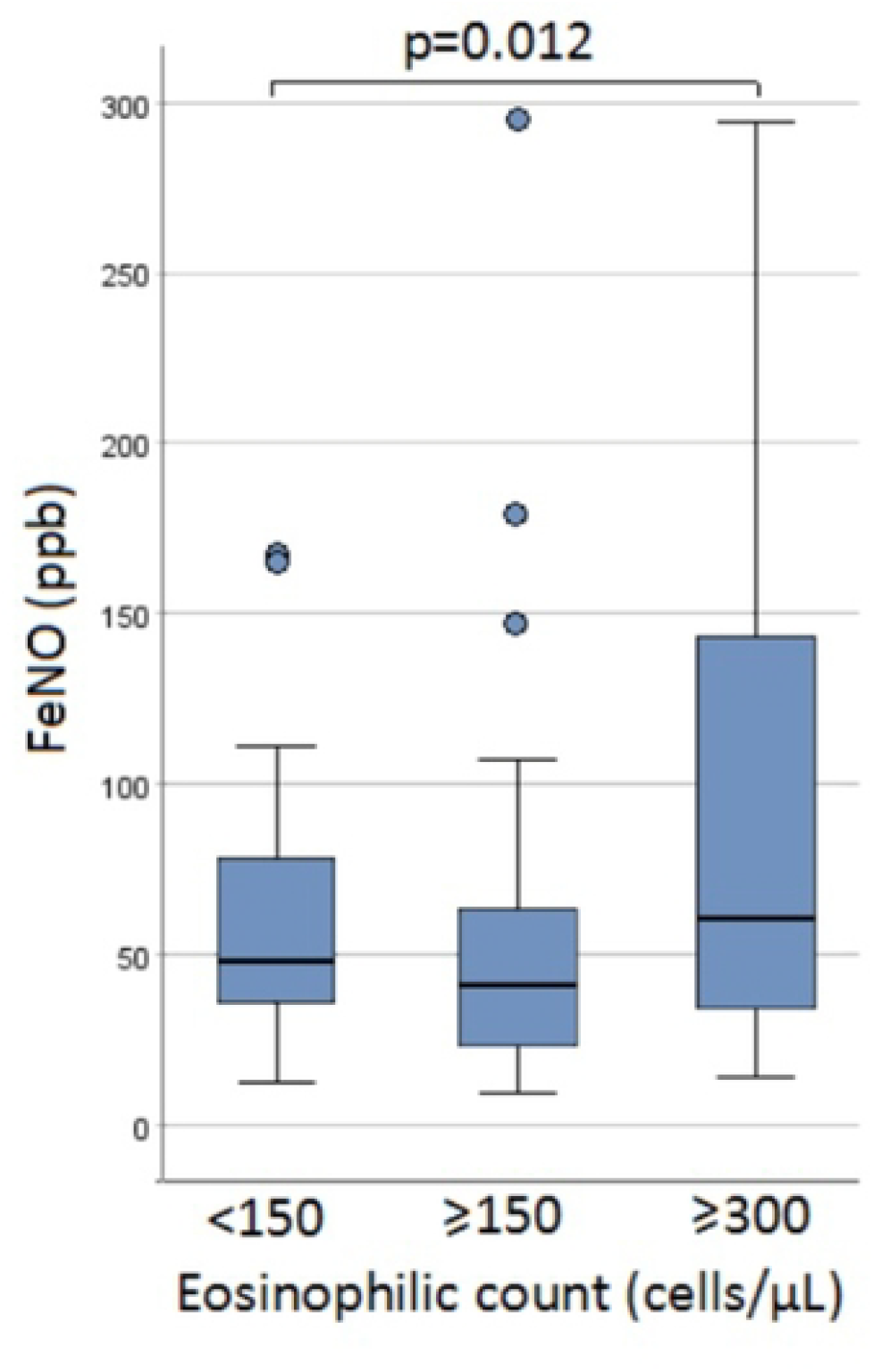
Fractional exhaled nitric oxide (FeNO) levels in relation to the absolute eosinophilic level in asthmatic patients.

**Figure 2:**
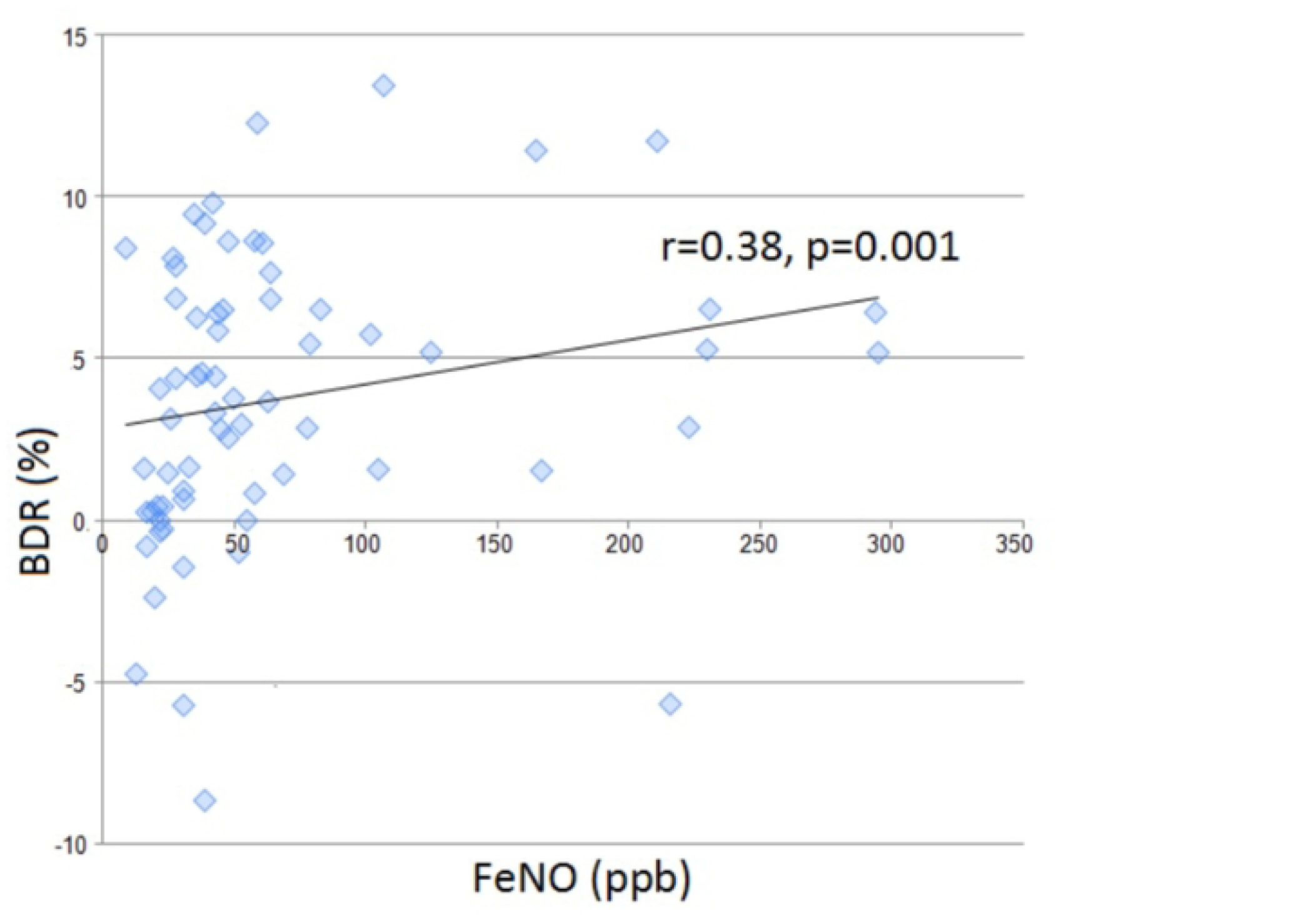
Correlation of fractional exhaled nitric oxide (FeNO) levels and bronchodilator responsiveness (BDR) in asthmatic patients.

The mean absolute peripheral eosinophils count was 310.5±344.2 cells/μL. 64 patients (63.4%) had peripheral eosinophils count of ≥150. Peripheral eosinophils tended to be higher in males (339.03±380.5 cells/μL vs 297.9±326.3 cells/μL, p=0.6), were comparable according to the smoking status (p=0.68), ACT status (p=0.11), presence of allergic rhinitis (p=0.78), chronic rhinosinusitis with nasal polyposis (p=0.1), GERD (p=0.71) and eczema/atopic dermatitis (p=0.12). The mean eosinophilic count was significantly higher in those with moderate airway obstruction assessed by FEV1% post-BD (641.8±548.4 cells/μL) compared to those with mild (277.8±295.5 cells/μL), moderately-severe (273.3±379 cells/μL, severe (180±42.4 cells/μL) and very severe grade in 1 case (160 cells/μL)(p=0.041)(Figure 3). The peripheral eosinophils did not significantly correlate with BMI (r=-0.07, p=0.48) or BDR (r=0.008, p=0.095). 14 (13.9%) cases had both high FeNO (>50 ppb) together with increased eosinophils >5%. The BDR was higher in those with elevated both biomarkers (5.5±5.3%) compared to those without (3.5±4.2%) (p=0.27).

**Figure 3:**
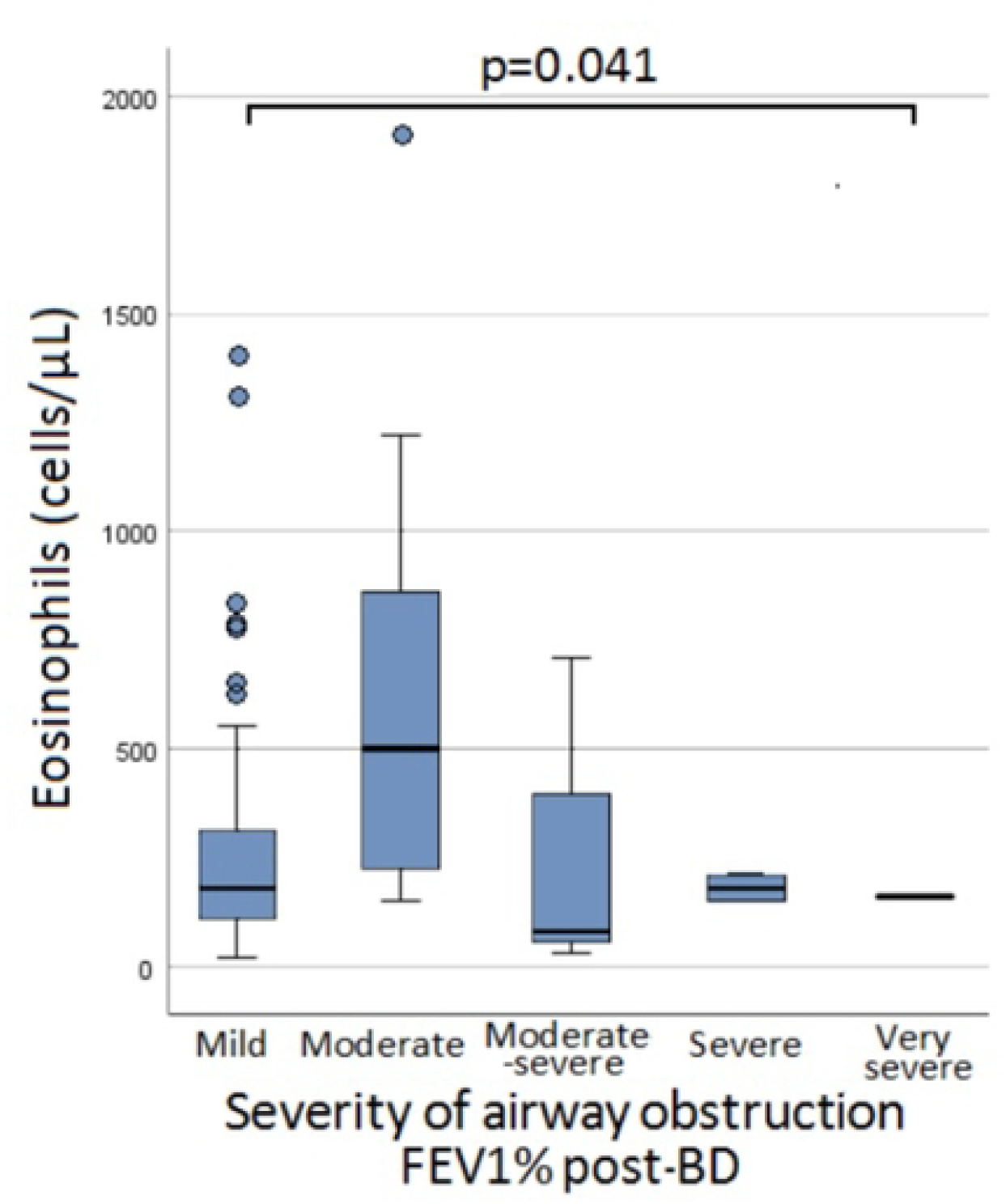
Eosinophilic count according to severity of airway obstruction assessed by forced expiratory volume 1 (FEV1%) post bronchodilatation (BD) in patients with asthma

There was a tendency of FeNO to discriminate patient with and without allergic rhinitis at a cut off of 32 ppb (p=0.059) with an AUC 0.61, 95%CI 0.5-0.72, sensitivity 79.3%, specificity 39.5% compared to eosinophils (p=0.6). Both eosinophils and FeNO similarly performed in differentiating chronic rhinosinusitis with nasal polyposis (p=0.17 and p=0.15 respectively) and poorly differentiated GERD (p=0.87 and p=0.85 respectively) and eczema/atopic dermatitis (p=0.11 and p=0.24 respectively) (Figure 4).

**Figure 4:**
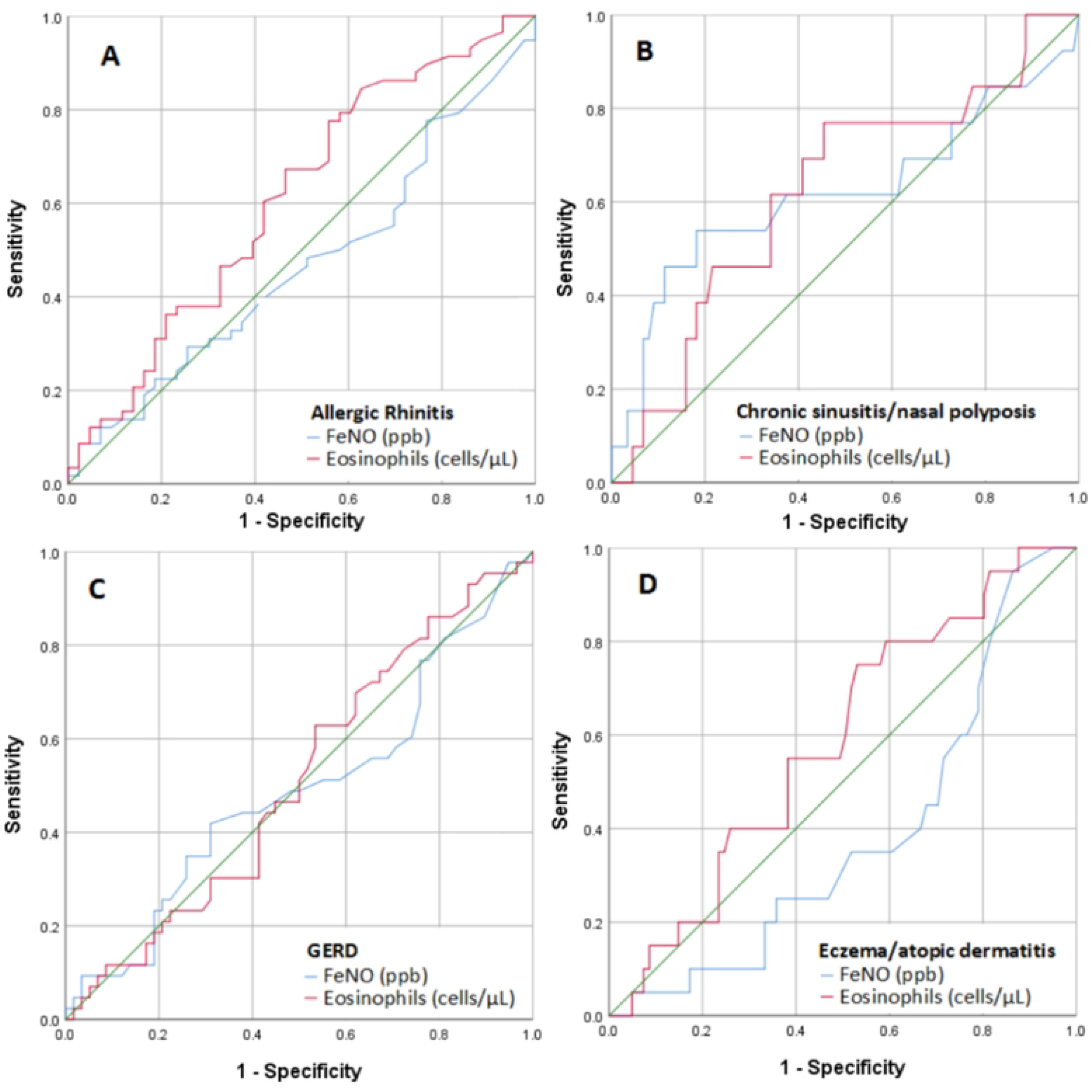
Receiver operating characteristic (ROC) curve for the ability of fractional exhaled nitric oxide (FeNO) and eosinophils to discriminate allergic rhinitis, chronic sinusitis with nasal polyposis, gastroesophageal reflux disease (GERD) and eczyma/atopic dermatitis in patients with asthma

## Discussion

This study explored the relationship between FeNO levels, peripheral eosinophil counts, and other clinical characteristics in a cohort of 101 adult asthmatic patients. The findings suggest a potential association between FeNO and type 2 inflammation, as evidenced by the significantly higher FeNO levels observed in patients with eosinophil counts ≥300 cells/µL compared to those with lower counts. The mean FeNO level was significantly higher in patients with peripheral eosinophil counts ≥300 cells/µL compared to those with lower eosinophils levels, consistent with existing literature demonstrating a strong association between FeNO and eosinophilic inflammation in asthma [11,18,19]. This is further supported by a growing body of evidence indicating that FeNO levels not only reflect eosinophilic inflammation but also predict responsiveness to anti-inflammatory therapies, serve as a reliable marker of type 2 airway inflammation in asthma, and underscore its potential role in identifying type 2 inflammation, a hallmark of eosinophilic asthma [12].

However, while FeNO tended to correlate with eosinophilic count (r=0.16, p=0.11), this correlation did not reach statistical significance, potentially due to the variability in eosinophilic levels influenced by factors such as corticosteroid use, environmental exposures, or comorbidities [17]. These results highlight the complementary, rather than interchangeable, roles of FeNO and peripheral eosinophilia in asthma evaluation. For instance, FeNO may be more useful in detecting active airway inflammation, particularly in type 2 inflammation, whereas peripheral eosinophil counts might provide better insights into systemic eosinophilic activity or inflammation related to comorbidities [20]. Clinical scenarios such as monitoring response to biologic therapies or adjusting inhaled corticosteroid doses could benefit from integrating both biomarkers to provide a more comprehensive assessment of asthma pathophysiology.

A significant positive correlation between FeNO levels and BDR (r=0.38, p=0.001) was observed, consistent with findings from previous studies such as Provenzano et al., which demonstrated FeNO’s potential as a predictive marker for bronchodilator response [21]. This highlights the utility of FeNO, particularly when integrated with other assessments, in refining asthma phenotyping and identifying patients who may benefit from targeted therapeutic strategies aligning with current clinical guidelines recommendation [18]. On the other hand, FeNO levels did not correlate with poor asthma control measured by ACT as previously known in the literature [18]. However, this can be attributed to poor asthma perception, a phenomenon well documented in the literature [22,23].

Despite the trend of elevation, our study found no significant correlation in FeNO levels across clinical characteristics of type 2 asthma, including allergic rhinitis, chronic rhinosinusitis, and eczema. This contrasts with the well-documented positive correlations between FeNO and these conditions in previous studies [24,25]. The lack of significance in our findings may be attributed to the limited sample size or the potential effects of corticosteroid treatment, which was not accounted for in our analysis. Further research with larger cohorts and detailed evaluation of treatment effects is warranted to better understand these relationships.

FeNO levels showed no significant differences across the various grades of airway obstruction as assessed by post-bronchodilator FEV1 (p = 0.81). However, higher FeNO levels were observed in patients with moderate obstruction compared to those with mild, severe, or very severe obstruction. This trend may reflect the active contribution of type 2 inflammation in moderate obstruction, while in severe or very severe obstruction, airway remodeling and structural changes dominate, leading to reduced FeNO levels despite ongoing disease. Additionally, patients with severe disease are more likely to be on corticosteroids or biologics, which suppress FeNO, whereas those with moderate obstruction may not require such intensive treatment, allowing FeNO to remain elevated. The limited sample size may also have skewed results, with a higher proportion of patients with active type 2 inflammation in the moderate obstruction group. These findings highlight FeNO’s role as a biomarker for inflammatory phenotypes of asthma, warranting further research with larger cohorts to validate these observations.

## Limitations

This study has limitations, including potential bias from its retrospective design, small sample size and the single-center setting, which may limit generalizability. The influence of treatment regimens, particularly corticosteroids, on FeNO and eosinophil levels was not fully accounted for, potentially confounding the results. Future prospective, multicenter studies are needed to validate these findings and explore the relationships between FeNO, eosinophils, and asthma phenotypes more comprehensively.

## Conclusion

This study demonstrates that FeNO is significantly associated with BDR and elevated peripheral eosinophil counts, underscoring its relevance as a marker of type 2 inflammation in asthma. Integrating FeNO with peripheral eosinophil measurements could provide a more comprehensive approach to asthma management by tailoring treatment strategies.

## Data Availability

All relevant data are within the manuscript and its supporting information files.

